# Association of heart rate variability and atrial fibrillation in patients with obstructive hypertrophic cardiomyopathy

**DOI:** 10.1101/2023.08.29.23294803

**Authors:** Changrong Nie, Zhengyang Lu, Changsheng Zhu, Minghu Xiao, Zining Wu, Qiulan Yang, Tao Lu, Yanhai Meng, Shuiyun Wang

## Abstract

**Background:** Atrial fibrillation (AF) is surprisingly common in patients with obstructive hypertrophic cardiomyopathy (oHCM) and is associated with significant symptoms and poor survival. Globally accepted models for AF detection are yet to be established. We aimed to investigate the relationship between heart rate variability (HRV) and AF in patients with oHCM.

**Methods:** We enrolled 1112 consecutively recruited patients with oHCM, including 158 and 954 patients with and without AF, respectively. The HRV variables mainly included the standard deviation of the mean R-R intervals (SDNN), root mean squared successive difference (rMSSD), and percentage of cycles differing from the preceding one by > 50 ms (pNN50). The SDNN, rMSSD, and pNN50 were transformed into binary variables underlying the cutoff for AF detection (termed SDNN_cutoff, rMSSD_cutoff, and pNN50_cutoff, respectively).

**Results:** The mean age of this cohort was 48.94 ± 12.37 years, and 451 patients were females. The patients with AF were older, more likely to have palpitations, had a larger left atrial diameter and lower left ventricular outflow gradient, and a significantly higher SDNN, rMSSD, and pNN50 than those without AF. In multivariable logistic regression analysis, pNN50_cutoff was independently associated with AF (odds ratio: 7.86, 95% confidence interval [CI]: 4.25–14.7), while the model including pNN50_cutoff had the largest area under the curve (0.736; 95% CI: 0.690–0.782) and the lowest Akaike information criterion (774).

**Conclusion:** HRV was associated with a higher incidence of AF. Among the HRV variables, pNN50_cutoff (cutoff value = 43.5) may be a good predictor of AF in patients with oHCM.

## Introduction

Atrial fibrillation (AF) is the most common arrhythmia in hypertrophic cardiomyopathy (HCM), occurring in approximately 20% of patients with HCM [1]. Patients with AF are more likely to present with symptoms, have a higher rate of hospitalization due to heart failure or thromboembolic events, and have a worse long-term prognosis [2, 3]. Given its high prevalence and morbidity, understanding the mechanisms and prediction of AF in patients with HCM are critical for its early prevention and intervention [4]. Multiple pathophysiological mechanisms underlying the development of AF have been studied in the general population [5, 6]. In recent years, increasing evidence has suggested that autonomic nervous system (ANS) dysfunction, including dysfunction of the sympathetic and parasympathetic nervous systems, is involved in the pathogenesis of AF [7, 8]. Previous studies confirmed the presence of ANS dysfunction in patients with HCM [9, 10]. However, no association between ANS dysfunction and AF has been reported in patients with HCM.

Heart rate variability (HRV) is the small change in time between consecutive cardiac cycles that can be used to quantitatively assess sympathetic and parasympathetic tone and balance in the cardiac ANS [11]. Noninvasive assessment of the cardiac autonomic status can be achieved using HRV analysis [12]. HRV has been extensively studied in the general population and has been shown to have an important prognostic impact on a variety of cardiovascular diseases [13–15]. Previous studies have suggested that ANS disorders are a potential trigger of AF onset and a factor in AF maintenance. Indeed, changes in ANS tension are known to play an important role in the occurrence, development, and persistence of AF [16]. Studies on the association between HRV and AF in patients with HCM are rare. Here, we retrospectively collected preoperative 24 h Holter monitoring HRV data from a large obstructive HCM (oHCM) population who underwent surgery and aimed to analyze the correlation between HRV and AF and to provide evidence for the prediction and early prevention of AF in patients with HCM.

## Methods

### Patient Population

Between January 2015 and December 2019, 1491 patients with obstructive HCM underwent septal myectomy at our institution. Of the 1491 patients, 379 were excluded for the following reasons: 1) absence of a preoperative ambulatory Holter electrocardiogram (ECG) evaluation, 2) prior history of heart surgery, and 3) preoperative permanent pacemaker installation. Consequently, 1112 patients were included in the final analysis. The diagnosis of HCM was based on the American Heart Association (AHA) and American College of Cardiology guidelines [17], which mainly included unexplained septal hypertrophy with a thickness > 15 mm or a septal cardium with a thickness > 13 mm, with a family history of HCM in the absence of other cardiac or systemic diseases. Septal myectomies were performed on patients whose symptoms were refractory to drug treatment and who had a left ventricular outflow tract (LVOT) gradient ≥ 50 mmHg at rest or during provocation. Septal myectomy (extended Morrow procedure) was performed as previously described [18]. All study patients signed informed consent forms before enrollment, and the study was approved by the Ethics Committee of Fuwai Hospital. All of the procedures were conducted in accordance with the ethical principles of the Declaration of Helsinki.

### Twenty-Four-Hour Holter ECG Monitoring

All patients in this study underwent 24-hour Holter electrocardiogram (ECG) monitoring (BI9800, Biomedical Instruments Co., Ltd., Osaka, Japan) 2-14 days prior to surgery. Sinus rhythm was mandatory for study entry, and each 24-hour Holter required at least 20 hours of artefact-free data. Holter recordings were computer analysed and all Holter readings were checked by the investigators. The time domain analysis of HRV included the standard deviation of normal-to-normal intervals (SDNN); the standard deviation of mean normal-to-normal intervals for each 5-minute segment of a 24-hour HRV recording (SDANN); the root mean square difference of successive normal-to-normal intervals (rMMSD); and the percentage of NN50 (normal-to-normal intervals >50 milliseconds) in the total number of RR intervals. In the time domain index, SDNN reflects total sympathetic and parasympathetic nervous system activity, while SDANN reflects sympathetic nervous system tone. RMSSD reflects changes in parasympathetic nervous system tone and pNN50 reflects parasympathetic nervous system tone activity.

### Echocardiography

Two experienced physicians conducted the echocardiographic examinations using an E9 ultrasound system (General Electric Company, Boston, MA, USA). All patients underwent preoperative and postoperative two-dimensional and Doppler echocardiography. Left atrial diameter (LAD), left ventricular (LV) end-diastolic diameter, LV ejection fraction, and ventricular septal thickness were measured according to the recommendations of the American Society of Echocardiography [19]. The LVOTG was calculated using the simplified Bernoulli equation. These methods are described in detail in our previous publication [18].

### AF Diagnosis and Evaluation

The diagnosis of AF met the definition of the 2020 European Society of Cardiology Guidelines [20]: A standard 12-lead ECG recording or a single-lead ECG tracing of ≥ 30 s showing a heart rhythm with no discernible repeating P waves and irregular RR intervals (when atrioventricular conduction is not impaired) is considered diagnostic of clinical AF. We collected data from a previous ECG, a 24 h Holter monitor, and the medical history to help diagnose AF before surgery.

### Statistical Analyses

Categorical variables are presented as numbers and percentages and continuous variables as means and standard deviations or medians with interquartile ranges, as appropriate. Differences in characteristics across groups were compared using ANOVA or the chi-square test. Correlations between continuous and categorical variables were assessed using Pearson’s or Spearman’s correlation coefficients, as appropriate. Receiver operating characteristic (ROC) curves were constructed to explore the optimal cutoff value of each HRV variable in detecting AF, before transforming HRV variables into categorical variables as candidates for the logistic regression analysis models. Univariate and multivariate logistic regression analyses were performed to determine the association between HRV and AF. All relevant clinical and echocardiographic variables were included in the multivariate model, with a p-value < 0.1. Multivariate logistic regression models were constructed by adjusting variables from the univariate analysis using a backward method, before adding each HRV variable to the multivariate logistic regression model separately as the final model. The Akaike information criterion (AIC) was used to compare the goodness of fit for each model, and model discrimination was assessed by comparing the area under the curve (AUC), net reclassification improvement (NRI), and integrated discrimination improvement (IDI). A nomogram was constructed based on the optimal model to estimate the probability of AF. All statistical analyses were performed using R version 4.2.2 (R Foundation for Statistical Computing, Vienna, Austria) and GraphPad Prism version 8.0 (GraphPad Software Inc., La Jolla, CA, USA).

## Results

### Clinical Characteristics of the Study Cohort

A total of 1112 consecutive patients with oHCM in this study, including 158 patients with AF and 954 patients without AF. The mean age of the study population was 48.94 ± 12.37 years and 451 (40.6%) patients were female. The baseline characteristics of those patients were grouped by patients with and without AF, and the results were summarized in Table 1. Compared to patients without AF, the patients with AF were older (52.53 ± 12.24 vs. 48.35 ± 12.30 years, p < 0.01), more likely to have palpitation (47.5% vs. 24.2%, p < 0.01), had a larger left atrial diameter (48.69 ± 7.09 vs. 44.49 ± 6.46 mm, p < 0.01), had a lower LVOT gradient (75.77 ± 28.89 vs. 84.80 ± 28.89 mmHg, p < 0.01). Moreover, SDNN (124.89±60.15 vs. 112.35±45.00, P=0.002), rMSSD (43.90±27.21 vs. 32.96±27.96, P<0.01), and pNN50 (19.91±22.33 vs. 10.75±11.95, P<0.01) were significantly increased in AF patients compared to those without AF. Other clinical variables in listed in Table 1 were comparable between the two groups.

**Table 1.**
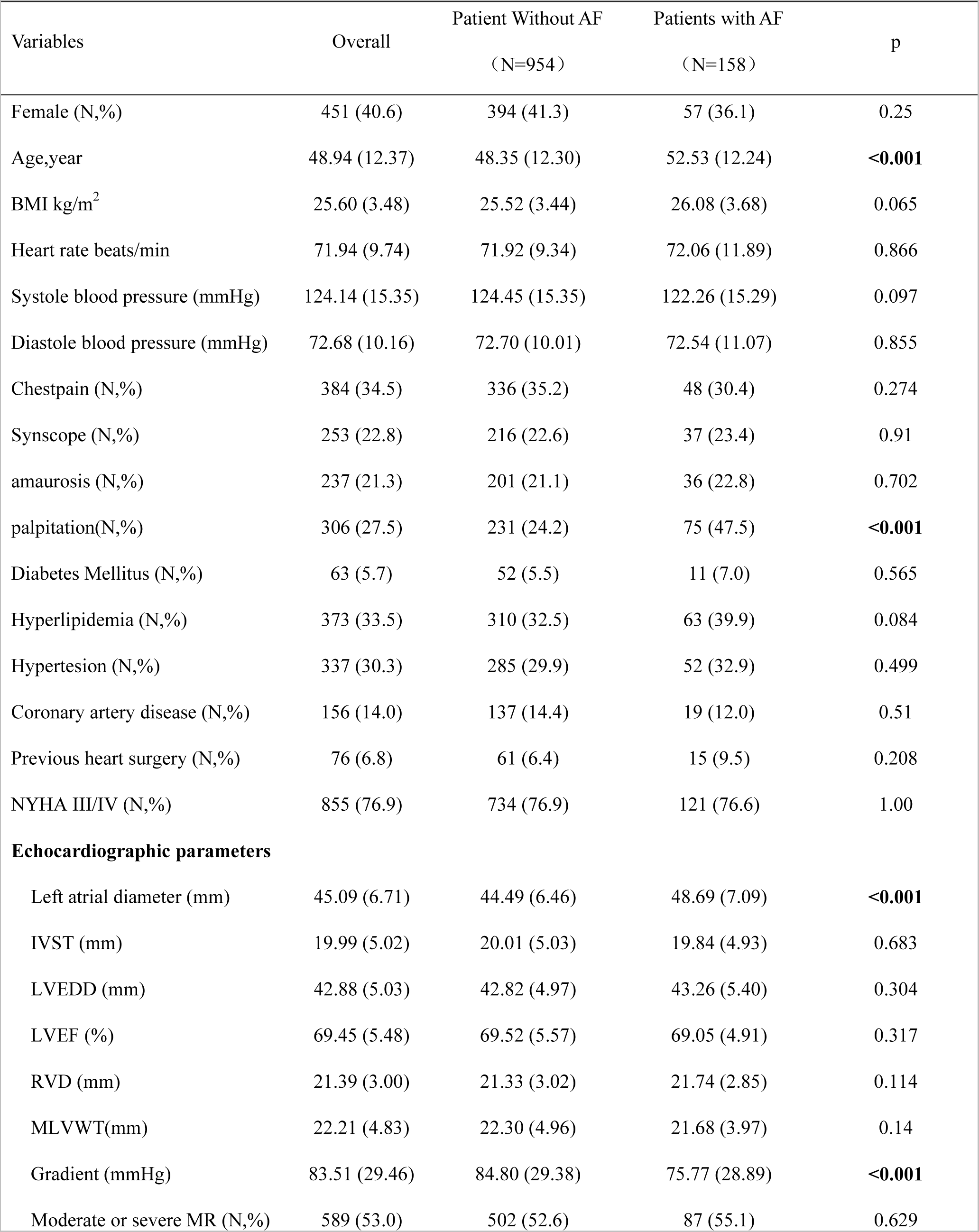
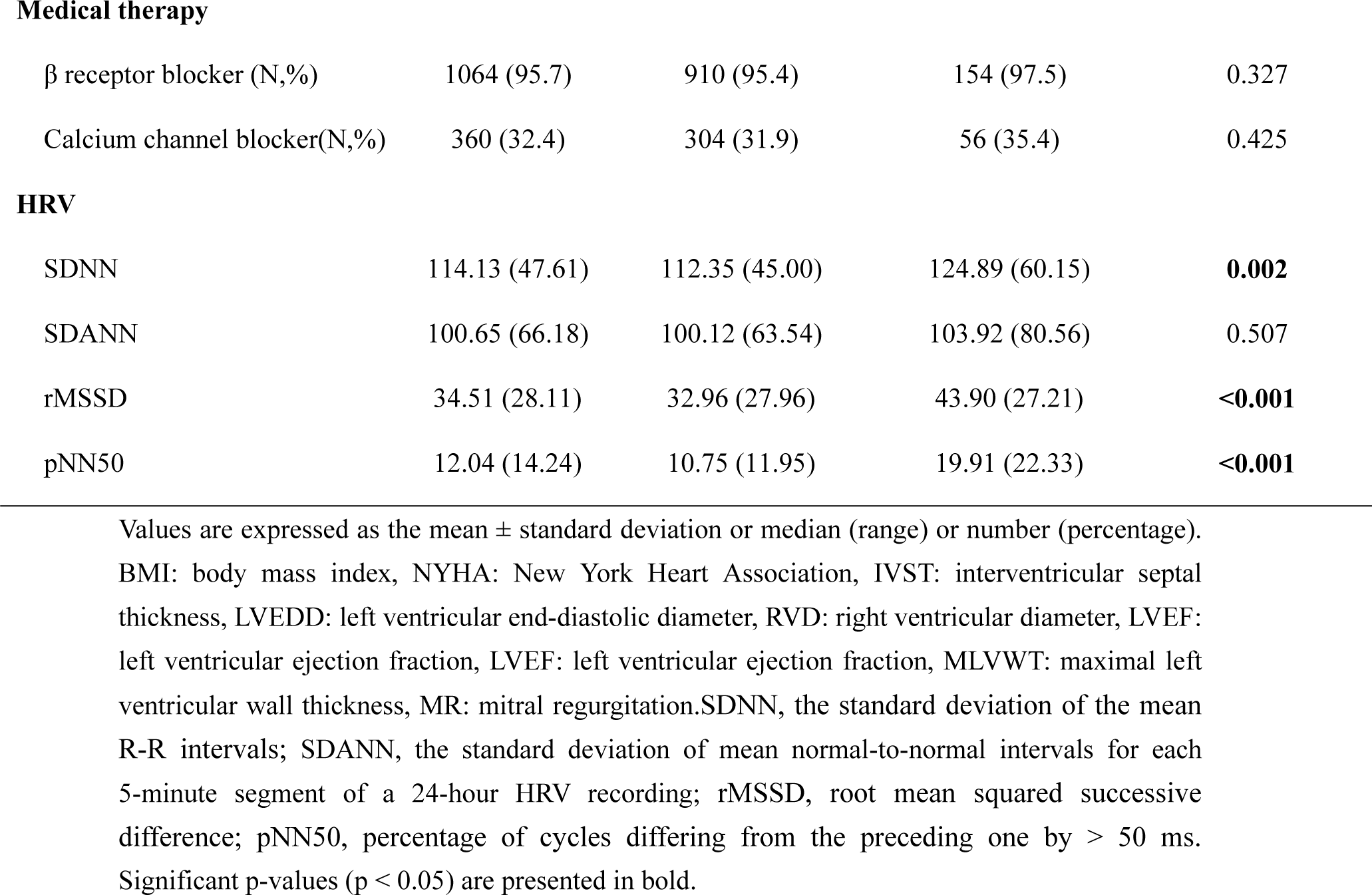
Baseline characteristics of the study population.

| Variables | Overall | Patient Without AF<br>(N=954) | Patients with AF<br>(N=158) | p |
| --- | --- | --- | --- | --- |
| Female (N,%) | 451 (40.6) | 394 (41.3) | 57 (36.1) | 0.25 |
| Age,year | 48.94 (12.37) | 48.35 (12.30) | 52.53 (12.24) | **<0.001** |
| BMI kg/m<sup>2</sup> | 25.60 (3.48) | 25.52 (3.44) | 26.08 (3.68) | 0.065 |
| Heart rate beats/min | 71.94 (9.74) | 71.92 (9.34) | 72.06 (11.89) | 0.866 |
| Systole blood pressure (mmHg) | 124.14 (15.35) | 124.45 (15.35) | 122.26 (15.29) | 0.097 |
| Diastole blood pressure (mmHg) | 72.68 (10.16) | 72.70 (10.01) | 72.54 (11.07) | 0.855 |
| Chestpain (N,%) | 384 (34.5) | 336 (35.2) | 48 (30.4) | 0.274 |
| Synscope (N,%) | 253 (22.8) | 216 (22.6) | 37 (23.4) | 0.91 |
| amaurosis (N,%) | 237 (21.3) | 201 (21.1) | 36 (22.8) | 0.702 |
| palpitation(N,%) | 306 (27.5) | 231 (24.2) | 75 (47.5) | **<0.001** |
| Diabetes Mellitus (N,%) | 63 (5.7) | 52 (5.5) | 11 (7.0) | 0.565 |
| Hyperlipidemia (N,%) | 373 (33.5) | 310 (32.5) | 63 (39.9) | 0.084 |
| Hypertesion (N,%) | 337 (30.3) | 285 (29.9) | 52 (32.9) | 0.499 |
| Coronary artery disease (N,%) | 156 (14.0) | 137 (14.4) | 19 (12.0) | 0.51 |
| Previous heart surgery (N,%) | 76 (6.8) | 61 (6.4) | 15 (9.5) | 0.208 |
| NYHA III/IV (N,%) | 855 (76.9) | 734 (76.9) | 121 (76.6) | 1.00 |
| **Echocardiographic parameters** |  |  |  |  |
| Left atrial diameter (mm) | 45.09 (6.71) | 44.49 (6.46) | 48.69 (7.09) | **<0.001** |
| IVST (mm) | 19.99 (5.02) | 20.01 (5.03) | 19.84 (4.93) | 0.683 |
| LVEDD (mm) | 42.88 (5.03) | 42.82 (4.97) | 43.26 (5.40) | 0.304 |
| LVEF (%) | 69.45 (5.48) | 69.52 (5.57) | 69.05 (4.91) | 0.317 |
| RVD (mm) | 21.39 (3.00) | 21.33 (3.02) | 21.74 (2.85) | 0.114 |
| MLVWT(mm) | 22.21 (4.83) | 22.30 (4.96) | 21.68 (3.97) | 0.14 |
| Gradient (mmHg) | 83.51 (29.46) | 84.80 (29.38) | 75.77 (28.89) | **<0.001** |
| Moderate or severe MR (N,%) | 589 (53.0) | 502 (52.6) | 87 (55.1) | 0.629 |
| Medical therapy |  |  |  |  |
| β receptor blocker (N,%) | 1064 (95.7) | 910 (95.4) | 154 (97.5) | 0.327 |
| Calcium channel blocker(N,%) | 360 (32.4) | 304 (31.9) | 56 (35.4) | 0.425 |
| HRV |  |  |  |  |
| SDNN | 114.13 (47.61) | 112.35 (45.00) | 124.89 (60.15) | **0.002** |
| SDANN | 100.65 (66.18) | 100.12 (63.54) | 103.92 (80.56) | 0.507 |
| rMSSD | 34.51 (28.11) | 32.96 (27.96) | 43.90 (27.21) | **<0.001** |
| pNN50 | 12.04 (14.24) | 10.75 (11.95) | 19.91 (22.33) | **<0.001** |

Values are expressed as the mean ± standard deviation or median (range) or number (percentage).
BMI: body mass index, NYHA: New York Heart Association, IVST: interventricular septal thickness, LVEDD: left ventricular end-diastolic diameter, RVD: right ventricular diameter, LVEF: left ventricular ejection fraction, LVEF: left ventricular ejection fraction, MLVWT: maximal left ventricular wall thickness, MR: mitral regurgitation. SDNN, the standard deviation of the mean R-R intervals; SDANN, the standard deviation of mean normal-to-normal intervals for each 5-minute segment of a 24-hour HRV recording; rMSSD, root mean squared successive difference; pNN50, percentage of cycles differing from the preceding one by > 50 ms. Significant p-values (p < 0.05) are presented in bold.

### Correlation Analysis Between HRV and Clinical Parameters

We next analyzed the correlation between the time-domain metrics of HRV and clinical data, and the results are summarized in Figure 1. The LAD positively correlated with rMSSD (r = 0.09, p < 0.01) and pNN50 (r = 0.089, p < 0.01). The preoperative LVOT gradients were negatively correlated with rMSSD (r = –0.10, p < 0.01) and pNN50 (r = –0.11, p < 0.01), while PNN50 was strongly correlated with rMSSD (r = 0.96, p < 0.01) and SDNN (r = 0.48, p < 0.01).

**Figure 1:**
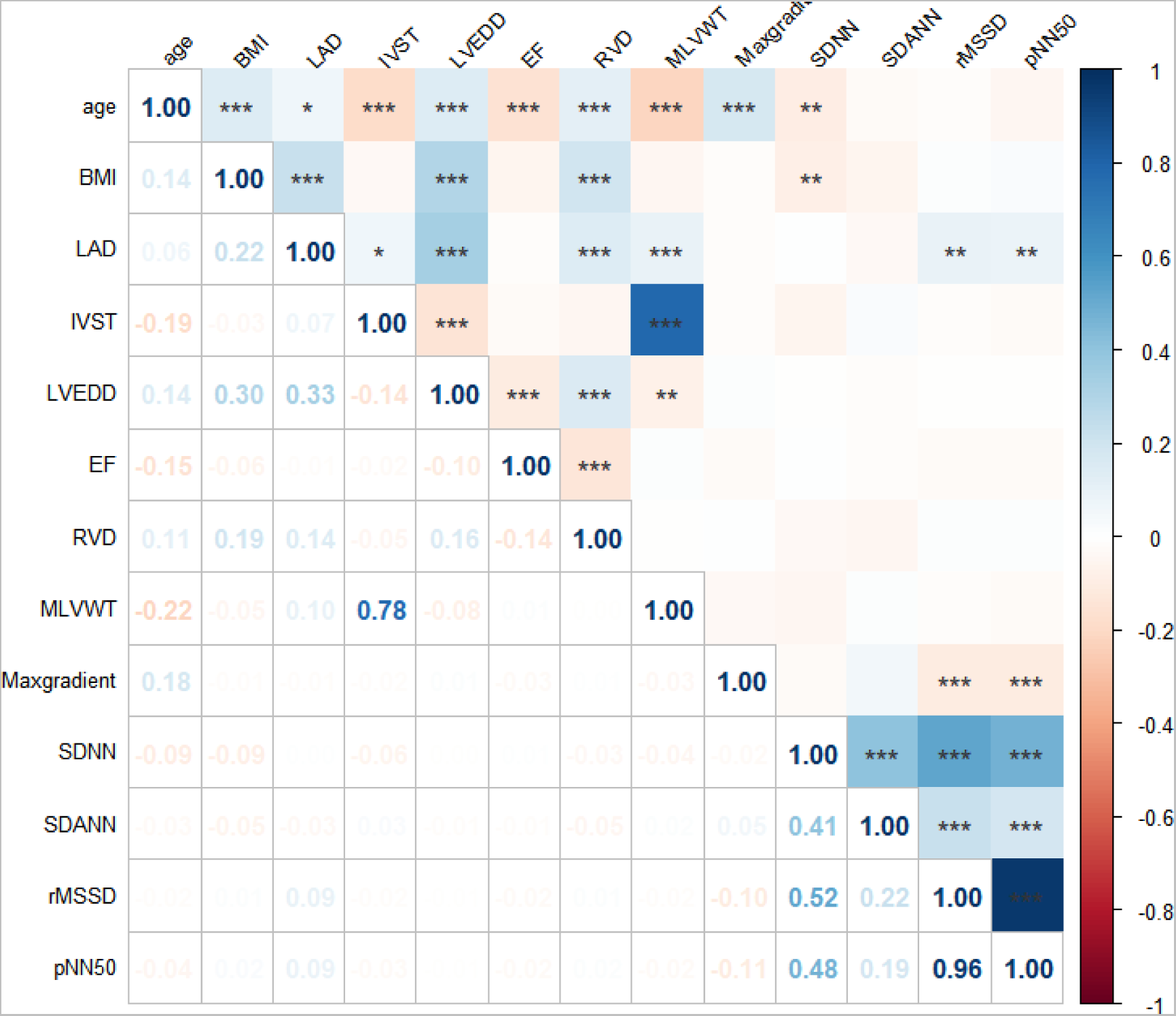
the correlation between the time-domain metrics of HRV and clinical data. BMI indicates body mass index; LAD, left atrial diameter; IVST, intraventricular septal thickness; EF, eject fraction; RVD, right ventricular diameter; MLVWT, maximal left ventricular wall thickness; maxgradient, maximal left ventricular outflow tract gradient; SDNN, the standard deviation of the mean R-R intervals; rMSSD, root mean squared successive difference; pNN50, percentage of cycles differing from the preceding one by > 50 ms.

### Univariate and Multivariate Regression Analyses of Risk Factors for AF Episodes

Before the univariate analysis, we plotted ROC curves to determine the optimal cutoff values of HRV variables in detecting AF (Figure 2). Next, we transformed the HRV variables into categorical variables as candidates for the logistic regression analysis models. The results are presented in Online Resource 2, and the cutoff values for the SDNN, rMSSD, and PNN50 were 153.5, 34.5, and 43.5, respectively. We transformed the SDNN, rMSSD, and PNN50 into new candidates (SDNN_cutoff, rMSSD_cutoff, and pNN50_cutoff) based on their cutoff values. The results of the univariate logistic regression analysis for each variable are summarized in Table 2. Age, body mass index (BMI), hyperlipidemia, LAD, and LVOT gradient were included in the multivariate regression analysis to construct the control model. The variables in the control model and HRV variables were used separately to construct new models. The results of the multivariate logistic regression analysis models for AF are presented in Table 3. After adjusting for age, BMI, hyperlipidemia, LAD, and LVOT gradients, SDNN, rMSSD, and pNN50 were independently associated with a higher incidence of AF, and SDNN_cutoff, rMSSD_cutoff, and pNN50_cutoff were independent indicators of AF. Among these variables, pNN50_cutoff had the largest effect size, with an odds ratio of 7.86 (95% confidence interval [CI]: 4.25–14.7).

**Figure 2:**
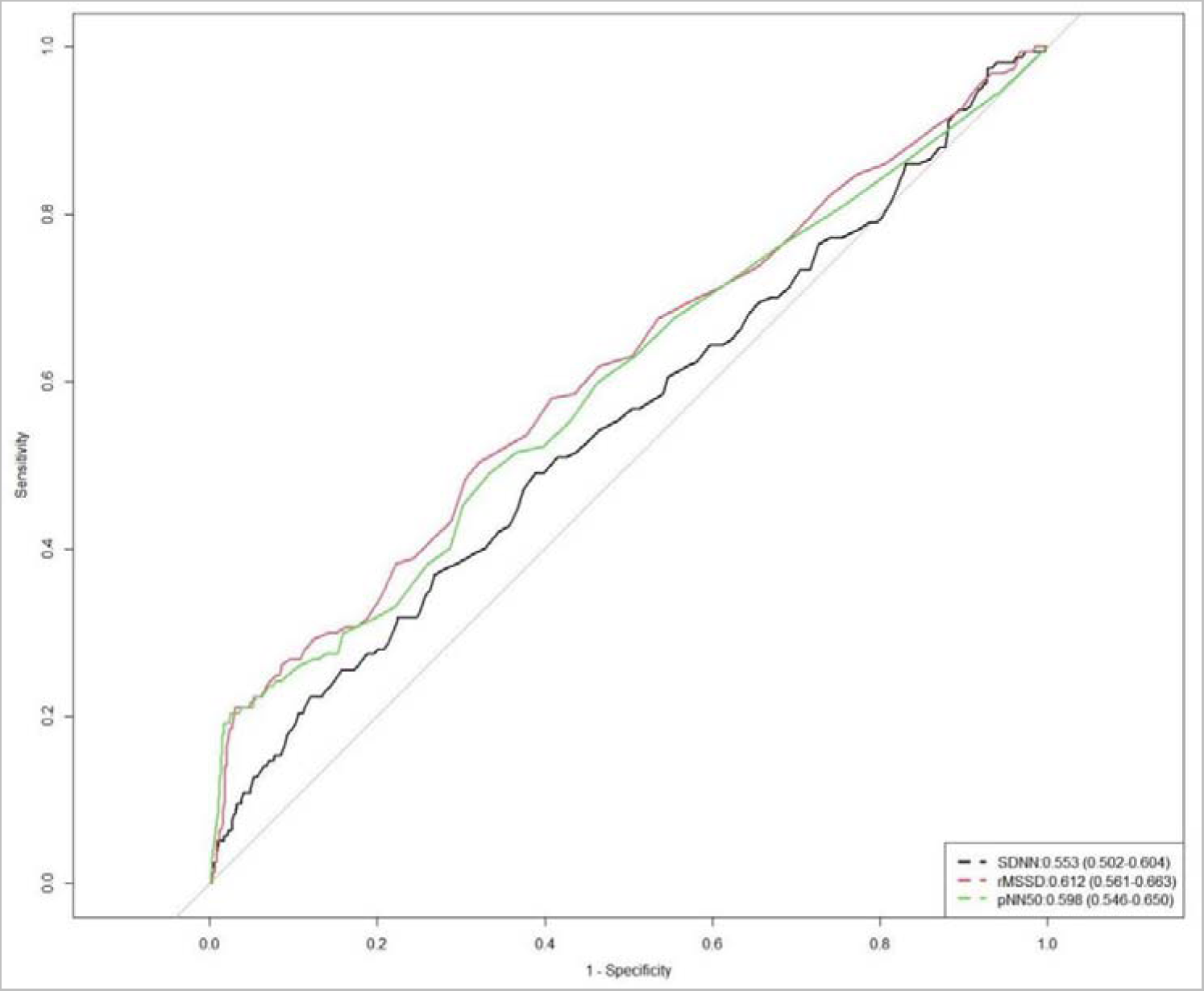
Receiver operating characteristic (ROC) curves were constructed to explore the optimal cutoff value of each HRV variable in detecting AF. The cutoff value of SDNN, rMSSD, pNN50 were 153.5, 34.5, and 43.5, respectively.

**Table 2.**
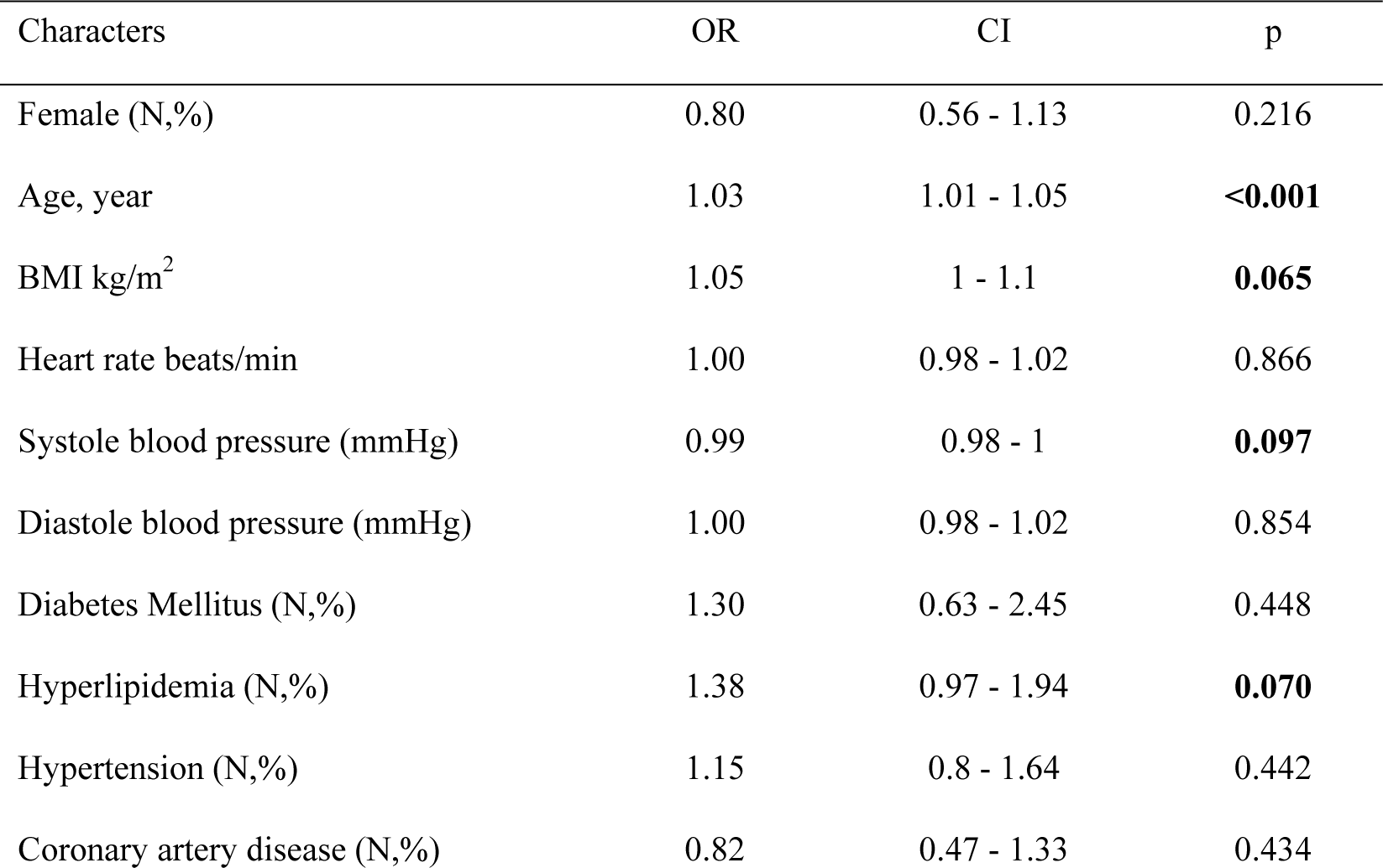
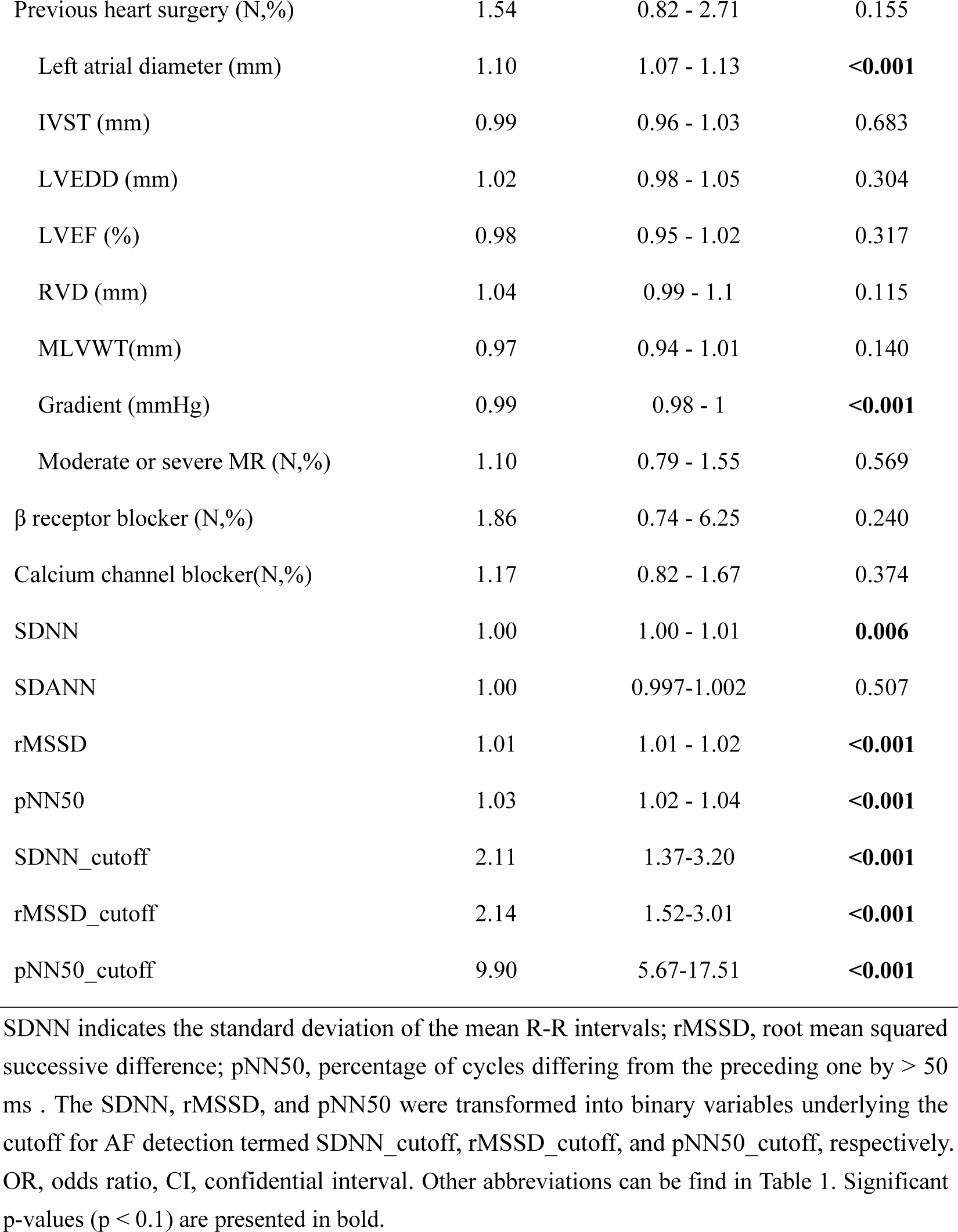
Univariate logistic regression analysis for AF.

**Table 3.**
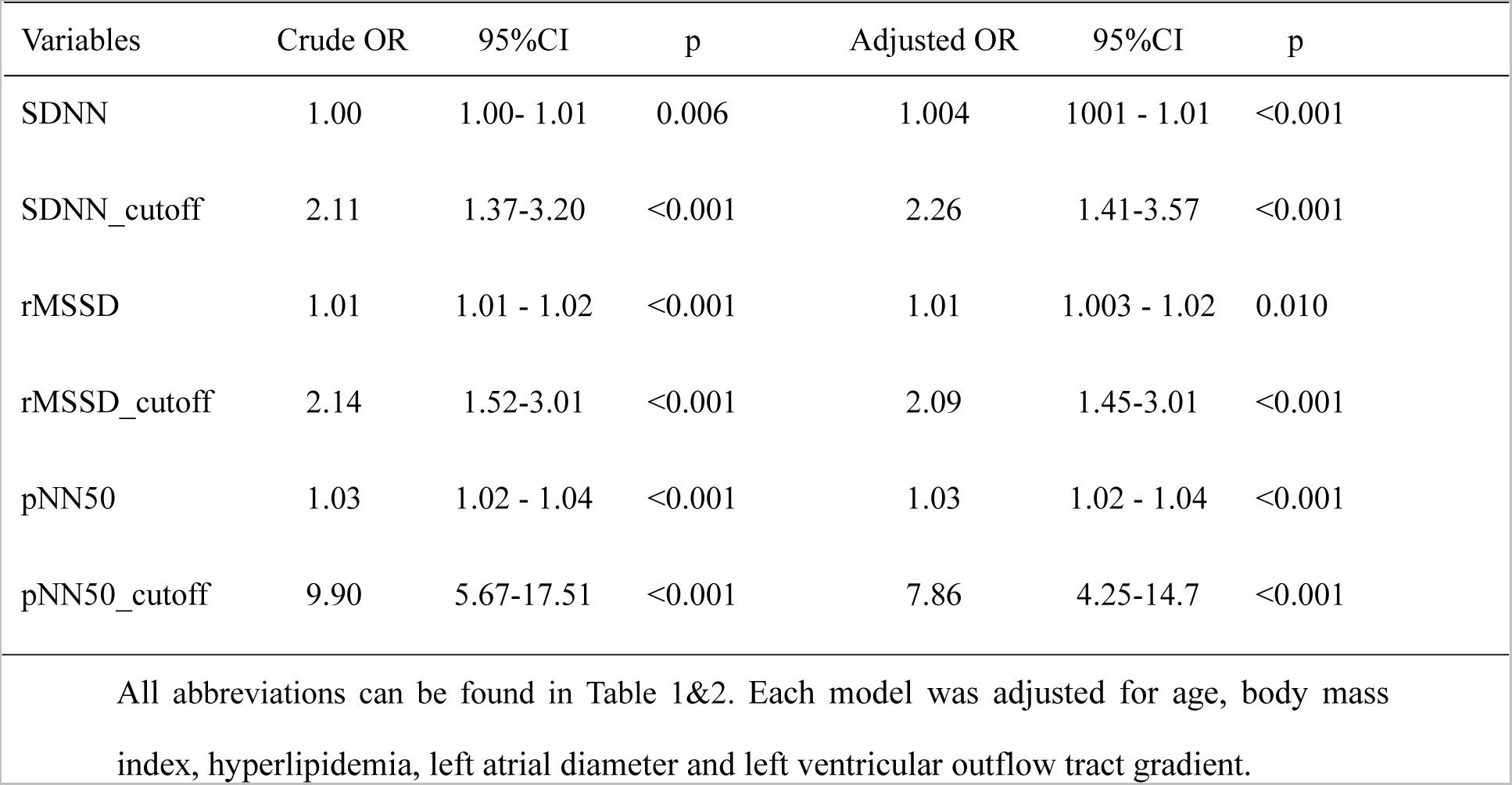
Multivariable logistic regression analysis models for AF.

| Variables | Crude OR | 95%CI | p | Adjusted OR | 95%CI | p |
| --- | --- | --- | --- | --- | --- | --- |
| SDNN | 1.00 | 1.00- 1.01 | 0.006 | 1.004 | 1.001 - 1.01 | <0.001 |
| SDNN_cutoff | 2.11 | 1.37-3.20 | <0.001 | 2.26 | 1.41-3.57 | <0.001 |
| rMSSD | 1.01 | 1.01 - 1.02 | <0.001 | 1.01 | 1.003 - 1.02 | 0.010 |
| rMSSD_cutoff | 2.14 | 1.52-3.01 | <0.001 | 2.09 | 1.45-3.01 | <0.001 |
| pNN50 | 1.03 | 1.02 - 1.04 | <0.001 | 1.03 | 1.02 - 1.04 | <0.001 |
| pNN50_cutoff | 9.90 | 5.67-17.51 | <0.001 | 7.86 | 4.25-14.7 | <0.001 |

### Assessing the Performance of HRV Models for Detecting AF

As mentioned above, we used the variables in the control model and HRV variables separately to construct new models. We then evaluated the performance of each new model, and the results are summarized in Table 4. Although the difference in AUC between the new models and the control model was not significant, the NRI and IDI in the new models improved significantly compared with those in the control model. The model including pNN50_cutoff had the largest AUC (0.736; 95% CI: 0.690–0.782) and the lowest AIC (774), indicating the best fit compared with the other models. A nomogram was developed based on the new pNN50_cutoff model to calculate the probability of incident AF (Figure 3A), including age, BMI, hyperlipidemia, LAD, LVOT gradient, and pNN50_cutoff. The calibration curve for the nomogram was close to the 45° diagonal for the most part, but slightly exceeded the diagonal at a high predicted risk, indicating that the nomogram is well calibrated in most situations but might underestimate the risk when the risk is already high (Figure 3B). The AUC of the new model of the pNN50_cutoff was 0.736 (95% CI: 0.690–0.782), demonstrating good discriminative ability of the model (Figure 3C).

**Figure 3:**
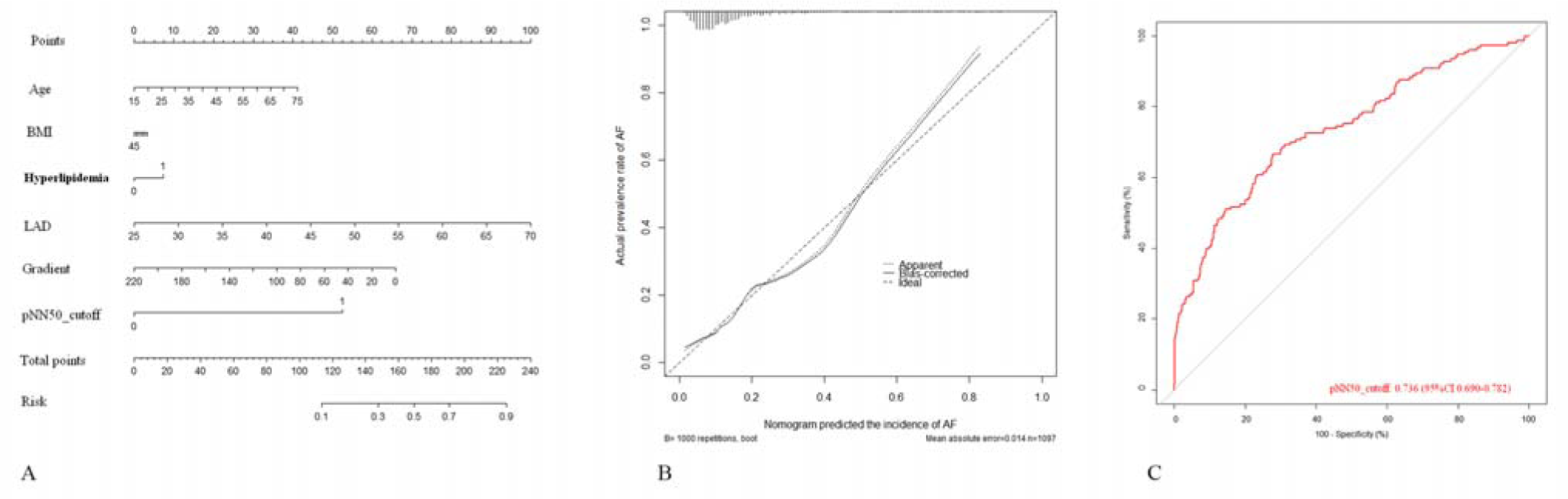
A, the nomogram was developed based on the pNN50_cutoff model to calculate the probability of incident AF, with predictors including age, BMI, hyperlipidemia, LAD, LVOT gradient and pNN50_cutoff; B, the calibration curve for the nomogram was mostly close to the 45°diagonal but slightly exceeded the diagonal at high predicted risk; C, the AUC of the new model of pNN50_cutoff was 0.736 (95% CI: 0.690-0.782).

**Table 4.** Performance of SDNN, rMSSD, and pNN50 for detecting atrial fibrillation.

|  |  |  |  |  |  |  |  |
| --- | --- | --- | --- | --- | --- | --- | --- |
| Variables | Discrimination |  | Reclassification |  |  |  | Goodness of fit |
|  | AUC (95%CI) | P | NRI (95%CI) | P | IDI (95%CI) | P | AIC |
| Control | 0.719<br>(0.673-0.765) | - | - | - | - | - | 813.7 |
| SDNN (vs.control) | 0.725<br>(0.679-0.770) | 0.367 | 0.264<br>(0.094-0.434) | 0.002 | 0.015<br>(0.004-0.026) | 0.006 | 804.8 |
| rMSSD (vs.control) | 0.731<br>(0.687-0.776) | 0.232 | 0.250<br>(0.083-0.417) | 0.003 | 0.041<br>(0.022-0.061) | <0.001 | 787.5 |
| pNN50 (vs.control) | 0.729<br>(0.684-0.774) | 0.367 | 0.238<br>(0.073-0.404) | 0.004 | 0.048<br>(0.026-0.070) | <0.001 | 784.0 |
| SDNN_cutoff (vs.control) | 0.721<br>(0.675-0.767) | 0.742 | 0.053<br>(-0.004-0.109) | 0.070 | 0.016<br>(0.006-0.026) | 0.002 | 804.6 |
| rMSSD_cutoff (vs.control) | 0.732<br>(0.687-0.776) | 0.195 | 0.070<br>(0.010-0.130) | 0.022 | 0.018<br>(0.008-0.030) | <0.001 | 800.1 |
| pNN50_cutoff (vs.control) | 0.736<br>(0.690-0.782) | 0.063 | 0.149<br>(0.074-0.225) | <0.001 | 0.064<br>(0.037-0.091) | <0.001 | 774.0 |

We further divided the SDNN, rMSSD, and pNN50 into five groups according to their quintile intervals and investigated the prevalence of AF in each group, as well as the prevalence of AF in patients having less than and more than the cut-off values of the SDNN, rMSSD, and pNN50. As shown in Figure 4, a positive Spearman’s linear association was observed between the incidence of AF and SDNN (Figure 4A), rMSSD (Figure 4B), and pNN50 (Figure 4C). The prevalence of AF in patients with SDNN > 153.5 or rMSSD > 34.5 (cutoff value) was 23.5% and 20.5%, respectively, nearly two-fold higher than those of their counterparts. Interestingly, we observed that the prevalence of AF in patients with pNN50 > 43.5 was 57.1%, five-fold higher than the values of their counterparts, which suggested that pNN50 was a predictor of AF with a high specificity in patients with oHCM.

**Figure 4:**
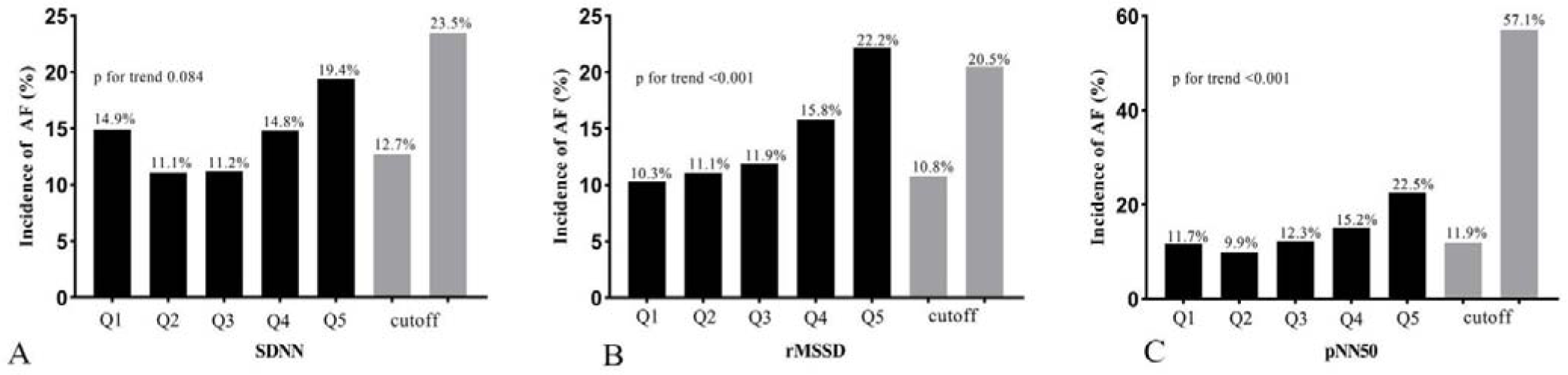
A positive Spearman’s linear association was observed between the incidence of AF and SDNN (Figure 4A), rMSSD (Figure 4B), and pNN50 (Figure 4C). The prevalence of AF in patients with SDNN > 153.5 or rMSSD > 34.5 (cutoff value) was 23.5% and 20.5%, respectively, the prevalence of AF in patients with pNN50 > 43.5 was 57.1%.

## Discussion

The findings of the current study can be summarised as follows. First, HRV variables were significantly increased in patients with AF compared to those without AF. SDNN, rMMSD and pNN50 were independently associated with a higher incidence of AF, and a positive Spearman’s linear association was observed between the incidence of AF and SDNN, rMMSD and pNN50. Second, in multivariable models, the model including pNN50_cutoff had the largest effect size, with an odds ratio of 7.86 (95% confidence interval [CI]: 4.25-14.7) for AF detection, while it had the largest area under the curve (0.736; 95% CI: 0.690-0.782) and the lowest Akaike information criterion (774). Third, in this study we constructed predictive models for AF in oHCM patients. The addition of HRV variables can improve the predictive ability of AF in oHCM patients, especially pNN50_cutoff.

The cardiac ANS contains both the extrinsic and intrinsic cardiac ANSs, including the sympathetic and parasympathetic nervous systems, the latter of which includes the epicardial autonomic ganglia, the fat pad, and the associated connecting nerve fibers, which essentially form a broad and highly connected epicardial neural network [21]. When activated, different physiological effects occur to maintain a stable balance and sinus rhythm in the heart. AF can be induced or promoted when the ANS is imbalanced [22]. In clinical practice, it is difficult to directly monitor autonomic nerve activity, and cardiac autonomic regulation and remodeling can only be evaluated indirectly. Therefore, considering the specificity and sensitivity of clinical surveillance, the selection of HRV parameters in a 24 h Holter monitor to reflect cardiac ANS activity is currently the primary research approach [12]. HRV describes the oscillations between the R-R intervals of a continuous heartbeat in Holter monitoring, which can be viewed as periodic changes in heart rhythm over time and constitutes a noninvasive method for qualitative and quantitative evaluation of the ANS. In this study, we analyzed the overall HRV profile of patients with HCM using a large sample of data. Our results demonstrated that the overall HRV of patients with HCM decreased compared with the normal values of each index, indicating that the activity of the ANS was reduced in patients with HCM. This is similar to the results of previous studies, most of which found that autonomic dysfunction in patients with HCM was dominated by decreased parasympathetic nervous system activity and increased sympathetic nervous system activity [23, 24]. It has also been established that autonomic dysfunction represented by reduced HRV is a risk factor for poor prognosis in cardiovascular diseases such as ventricular arrhythmias and sudden death [25]; however, to the best of our knowledge, no previous study has conducted HRV analysis of AF in patients with HCM.

AF occurs via various mechanisms, including electrical, structural, and neural remodeling, inflammation, and oxidative stress [5, 6]. In recent years, scholars have found that the cardiac ANS plays a significant role in the development of AF [7]. We demonstrated that the SDNN (representing total autonomic activity), and RMSSD and PNN50 (representing parasympathetic activity), were significantly increased in patients with HCM who developed AF compared with those of patients who did not, suggesting that total ANS activity and parasympathetic nervous system activity are increased in patients with HCM. We speculate that patients with HCM who have experienced AF have increased autonomic activity predominantly based on increased parasympathetic activity, in addition to the original ANS tone, which has been previously shown to be of great significance in the development and maintenance of AF [26]. Acetylcholine released in the physiological state of the parasympathetic nerves binds to M_2_ receptors on cardiomyocytes, causing a decrease in rhythm, conduction, and myocardial contractility, thereby inhibiting cardiac activity, and thus, parasympathetic nerves weaken the heart under normal conditions [27]. Smeets et al. [28] demonstrated that moderate stimulation of the parasympathetic nervous system shortened the impulse length and decreased the size of reentrant circuits, contributing to AF. Moreover, by stimulating the bilateral cervical vagus nerves, Schauerte et al. [29] reduced the effective refractory period at various atrial sites, triggering AF. This evidence suggests that the ANS activation can trigger AF by causing changes in atrial electrophysiology through acetylcholine release by nerve endings. AF also interferes with the distribution and function of the vagus nerve in the atria, which increases vagus tension and stabilizes the AF. A recent Mendelian randomization study [30] confirmed a significant correlation between HRV measurements and new-onset AF in the general population, supporting a causal relationship between the two. Furthermore, Fioranelli et al. [31] observed a decrease in LF and LF/HF and an increase in HF in patients with paroxysmal AF without structural heart disease, suggesting an elevated vagal tone, which is consistent with our findings.

Our results demonstrated that the SDNN, rMSSD, and pNN50 all correlated with AF occurrence in patients with HCM and were independent predictors of AF. Moreover, adding each of the three to the AF prediction model improved the predictive ability, especially pNN50 and rMSSD, further confirming the relationship between changes in parasympathetic tone and AF occurrence in patients with HCM. This is similar to the findings that the rMSSD and PNN50 are independent risk factors for AF recurrence after radiofrequency ablation, in which parasympathetic nerves are believed to play a key role [16]. The results of a recent meta-analysis of HRV similarly confirmed previous findings [32], in that the higher the SDNN and rMSSD, the more imbalanced the autonomic regulation and the predominance of parasympathetic regulation and the more easily AF is maintained. Previous studies have demonstrated that sympathetic excitation is associated with the development of ventricular arrhythmias [33] and that when sympathetic excitability is relatively enhanced, myocardial electrical stability is diminished, which predisposes individuals to the induction of malignant ventricular arrhythmias. In contrast, our results demonstrated that increased parasympathetic activity was associated with the development of AF in patients with HCM.

We also found a negative correlation between the SDNN and both age (p < 0.01) and BMI (p < 0.01), suggesting that total autonomic tone activity decreases with age and BMI. The results of our previous study demonstrated that age and BMI are independent risk factors for perioperative AF in patients with HCM [34], suggesting that HRV may play a role in the development of AF through age and BMI. Moreover, LAD was positively correlated with the rMSSD (p < 0.01) and pNN50 (p < 0.01), although the correlation between left atrial size and AF has been widely verified [35], suggesting that changes in parasympathetic nervous system tension may be associated with the occurrence of AF through changes in left atrial size. Furthermore, the preoperative LVOT gradient was negatively correlated with the rMSSD (r = –0.10, p < 0.01) and pNN50 (r = –0.11, p < 0.01), which suggests that for patients with HCM, the higher the LVOT gradient, the lower the parasympathetic tone and the less prone they will be to AF. Previous findings in HCM have suggested that AF is less likely to occur in patients with oHCM [36]. Our study provides an alternative explanation for these results, by showing that the LVOT gradient in patients with HCM may influence parasympathetic tone, which in turn influences the occurrence of AF.

Some articles [32, 37] have reported ANS changes before and after paroxysmal AF episodes, with the findings suggesting that a sympathetic and parasympathetic imbalance plays an important role in AF episodes; however, we did not analyze HRV before and after AF episodes in 24 h Holter results. Instead, we recorded the overall HRV changes and excluded patients whose 24 h Holter results included AF episodes. Prolonged HRV measurements (24 h) are more appropriate for estimating an individual’s basal autonomic state. Short-term (5 min) HRV measurements provide information about the system dynamics that disrupt basal homeostasis and may be a part of the trigger for arrhythmias as well as atrial or ventricular premature beats [38].

Our study has some limitations that warrant discussion. First, our outcome indicators were not completely accurate because the diagnosis of AF was derived from the patients’ ECG results, which may have missed some patients who had AF episodes and did not undergo electrocardiography. In addition to choosing objective indicators such as ECG parameters, we further determined the outcome indicators based on the patient’s medical history and description or clear diagnosis from other hospitals and medication history (e.g., amiodarone and ibutilide). Second, for technical reasons, it was necessary to exclude patients with multiple atrial premature beats during the period of ambulatory ECG recordings, who may have been the target population for our study. Third, as this was a retrospective study, no causal conclusions could be drawn. Fourth, many patients received medication during the 24 h Holter recording period, which may have affected HRV measurements. However, at baseline, the proportion of medications used in patients with and without AF did not differ significantly, suggesting that the medication itself had no significant effect. HRV is only an indirect measure of cardiac autonomic tone; therefore, the results of the present study should be interpreted with caution.

## Conclusion

HRV variables were significantly increased in patients with AF compared to those without AF. SDNN, rMMSD and pNN50 were independently associated with a higher incidence of AF, and a positive Spearman’s linear association was observed between the incidence of AF and SDNN, rMMSD and pNN50. The addition of HRV variables can improve the predictive ability of AF in oHCM patients, especially pNN50_cutoff. Among the HRV variables, pNN50_cutoff (cutoff value = 43.5) may be a good predictor of AF in patients with oHCM.

## Statements and Declarations

### Ethics approval

All procedures performed in studies involving human participants were in accordance with the ethical standards of the institutional and/or national research committee and with the 1964 Helsinki Declaration and its later amendments or comparable ethical standards. The study was approved by the Ethics Committee of Fuwai Hospital.

### Consent to Participate

Informed consent was obtained from all individual participants included in the study.

### Consent to Publish

Patients signed informed consent regarding publishing their data.

### Data, material, and/or code availability

The datasets generated and/or analysed in the current study are not publicly available due to Fuwai Hospital system, but data can be obtained from the corresponding author under reasonable request and with the permission of the Ethics Committee of Fuwai Hospital.

## Acknowledgments

The authors’ deepest gratitude goes to the Information Center of Fuwai Hospital for providing us data collection of this study.

## Non-standard abbreviations and acronyms

AF: Atrial fibrillation
BMI: Body mass index
LAD: Left atrial diameter
oHCM: Obstructive hypertrophic cardiomyopathy
HRV: Heart rate variability

